# Validation of an AI-Assisted Framework for Systematic Bias Assessment in Observational Studies

**DOI:** 10.64898/2026.04.26.26351778

**Authors:** Mahyar Etminan, Ramin Rezaeianzadeh, Antonios Douros

## Abstract

**Background:** The rapid expansion of medical literature has led to substantial variability and frequent contradictions in study findings, making it increasingly difficult to distinguish meaningful signals from noise. Much of this variability arises from differences in study methodology, where biases such as confounding, selection bias, and reverse causation can drive spurious associations. While artificial intelligence (AI)-assisted tools have been developed to support risk-of-bias assessment, most are designed for systematic reviews and are not tailored to identifying specific epidemiologic biases in observational studies. This highlights the need for structured, scalable approaches to evaluate study validity in real-world evidence.

**Objective:** To develop and validate an AI-assisted, expert-informed, rule-based framework (EpiVise) for systematically identifying and classifying key sources of bias in pharmacoepidemiologic studies, and to assess its agreement with expert evaluation.

**Methods:** We conducted a validation study using recently published pharmacoepidemiologic studies from high-impact journals (post-2025). Each study was independently assessed by the framework and two expert epidemiologists, across predefined bias domains, including measured confounding, confounding by indication, selection bias, immortal time bias, and disease latency. Agreement was evaluated using weighted kappa statistics. In the absence of a gold standard, expert judgment served as the reference benchmark. In a second phase, synthetic study scenarios with predefined embedded biases were constructed to assess the framework’s ability to detect known bias structures under controlled conditions.

**Results:** In analyses of published studies (10 studies; 60 ratings), agreement between the framework and expert assessments was substantial (κ = 0.75; 95% confidence interval [CI], 0.60–0.86), with 12 discordant ratings (20.0%), all limited to adjacent categories and occurring primarily in the confounding by indication and selection bias domains. In synthetic study scenarios (10 studies; 50 ratings), agreement was similarly substantial, with 42 of 50 ratings concordant (84%) and a weighted kappa of 0.77 (95% CI, 0.67-0.87); discordances included both adjacent-category and extreme disagreements and were concentrated in confounding by indication, selection bias, and prevalent user bias domains.

**Conclusions:** This AI-assisted, expert-informed framework, EpiVise provides a scalable and reproducible approach for evaluating epidemiologic study validity, substantial demonstrating agreement comparable to expert assessment. By systematically identifying key sources of bias, the framework has the potential to enhance the rigor and consistency of evidence evaluation, support peer review, and inform clinical, regulatory, and policy decision-making. Further validation across broader study designs and domains is warranted.

## INTRODUCTION

The rapid expansion of medical literature, with studies published daily, has led to substantial variability in findings, making it increasingly difficult to distinguish meaningful signals from noise. This variability can result in confusion among patients around the world, as well as clinicians, policymakers, and other stakeholders. For example, a recent systematic review reported an association between acetaminophen use and increased risk of autism^1^, generating concern among parents of young children worldwide, although subsequent analyses have questioned the robustness of this association.^2^ Similarly, individual studies may produce conflicting findings in rapid succession, for instance, one study may suggest that vitamin D is beneficial for a given disease, only to be contradicted by another study published shortly thereafter, highlighting the challenges in interpreting observational evidence.^3,4^

Much of this variability stems from differences in study methodology, leading to the coexistence of both high- and low-quality evidence. For instance, observational studies over many years suggested that statin use was associated with a reduced risk of Alzheimer’s disease; however, this protective effect was refuted in randomized controlled trials.^5^ Such discrepancies highlight how methodological biases, rather than true causal effects, can drive study findings. Identifying key sources of bias, including confounding, selection bias, and reverse causation, requires specialized expertise and systematic approaches to evaluate the validity of observational studies. ^6,7^

Prior studies have demonstrated that artificial intelligence (AI)-assisted tools can support risk-of-bias assessment by improving efficiency, consistency, and standardization in evidence synthesis, with performance approaching that of human reviewers in selected domains.^8,9^ However, most existing platforms are primarily designed and validated for systematic reviews and are not tailored to identifying and classifying specific epidemiologic biases in observational studies. As a result, their applicability to complex, real-world evidence remains limited. This highlights the need for structured, expert-informed approaches specifically developed to evaluate bias in observational research.

We created the first AI-assisted, expert-informed, rule-based framework, EpiVise to systematically assess epidemiologic studies by identifying key sources of bias and methodological limitations using a structured, rule-based framework grounded in established epidemiologic principles. By applying standardized criteria across studies, the framework has the potential to enhance the consistency and rigor of bias assessment, support peer review, and improve the overall quality and interpretation of observational research.

## METHODS

The framework (EpiVise, version 1.0) is an AI-assisted, expert-informed framework developed to systematically evaluate methodological quality and identify key sources of bias in observational studies. EpiVise applies structured, rule-based criteria grounded in established epidemiologic principles^10,11^, to ensure consistent and reproducible assessment. The framework was operationalized using a large language model (ChatGPT, OpenAI) to apply a structured, rule-based approach for extracting and evaluating key methodological features from each study. Six bias domains were defined a priori based on established epidemiologic principles: (1) confounding, (2) confounding by indication, (3) selection bias, (4) immortal time bias, (5) disease latency, and (6) prevalent user bias. The definition of the domains were defined and agreed up on by two experts. Each domain was assessed using standardized criteria and classified on a three-level scale (0 = unlikely, 1 = possible, 2 = likely), reflecting the strength of evidence for a given bias mechanism. The underlying decision rules were developed independently of the language model, remained fixed throughout the evaluation, and were applied consistently across all studies; detailed operational specifications are proprietary.

To enhance independence of validation and minimize potential overlap with information available during model development, we evaluated the framework using recently published pharmacoepidemiologic studies from higher-impact journals including Jama Network Open, JAMA Internal Medicine and Pharmacoepidemiology and Drug Safety restricting inclusion to those published after July 2025.

We applied EpiVise 1.0 to two sets of studies. First, we selected 10 pharmacoepidemiologic studies ^12–21^ published after July 2025. Two expert pharmacoepidemiologists (ME, AD) independently reviewed and scored each study using a three-level scale (0, 1, 2), with discrepancies resolved by consensus. The same studies were then evaluated using the framework, and agreement between expert and the framework assessments was quantified using a weighted kappa statistic.

In the second phase, we developed 10 fictitious study methodologies in which the same bias domains evaluated in the first phase were systematically embedded. Measured confounding was not assessed because these scenarios did not contain real patient data or covariate structures. Agreement between EpiVise and expert ratings was then evaluated using weighted kappa.

These scenarios were evaluated using the same scoring framework, and agreement was again assessed using a weighted kappa. Since this step did not include real published studies, the variable ‘confounding’ was not included as one of the assessed biases.

## RESULTS

For the analysis of published studies, 10 studies were evaluated across six bias domains, yielding 60 comparable bias ratings. Twelve ratings (20.0%) were discordant, with disagreements occurring most frequently in the selection bias (n=4) and confounding by indication (n=3) domains (Table 1). Additional discordances were observed in disease latency (n=2), prevalent user bias (n=2), and immortal time bias (n=1). All discordances were limited to adjacent categories (0 vs 1 or 1 vs 2), with no extreme disagreements. Overall agreement was substantial, with a weighted kappa of approximately 0.75 (95% CI, 0.60-0.86).

**Table 1.** Comparison of Bias Assessments Between an AI-Assisted Framework and Expert Evaluation in Published Studies.

| Study | Confounding |  | CBI* |  | Selection Bias |  | ITB** |  | Disease Latency |  | PUB+ |  |
| --- | --- | --- | --- | --- | --- | --- | --- | --- | --- | --- | --- | --- |
|  | Expert | AI | Expert | AI | Expert | AI | Expert | AI | Expert | AI | Expert | AI |
| **Algabani** | 0 | 0 | 1 | 1 | 1 | 1 | 0 | 0 | 1 | 1 | 1 | 1 |
| **De Burgos-González** | 0 | 0 | 1 | 1 | 0 | 0 | 0 | 0 | 0 | 1 | 1 | 0 |
| **Dumas** | 1 | 1 | 0 | 0 | 0 | 1 | 0 | 0 | 1 | 1 | 0 | 0 |
| **Hsiao** | 1 | 1 | 0 | 0 | 2 | 2 | 2 | 1 | 0 | 0 | 2 | 1 |
| **Leung** | 0 | 0 | 0 | 0 | 0 | 0 | 0 | 0 | 0 | 0 | 2 | 2 |
| **Lin** | 0 | 0 | 0 | 1 | 2 | 1 | 0 | 0 | 2 | 2 | 1 | 1 |
| **Moslemzadeh** | 0 | 0 | 1 | 0 | 0 | 0 | 0 | 0 | 0 | 1 | 0 | 0 |
| **Taniguchi** | 0 | 0 | 0 | 0 | 1 | 1 | 0 | 0 | 0 | 0 | 0 | 0 |
| **Wang** | 0 | 0 | 0 | 1 | 0 | 1 | 0 | 0 | 0 | 0 | 1 | 1 |
| **Portella** | 0 | 0 | 0 | 0 | 0 | 1 | 0 | 0 | 0 | 0 | 0 | 0 |

\*= Confounding by indication
\*\*= Immortal time bias
+ = Prevalent user bias
Highlighting indicates discordant scores; shaded cells represent differences between expert and AI platform assessments (Expert = orange, AI platform = pink).

Across the 10 fictitious studies and 5 bias domains (50 total ratings), agreement between expert and framework assessments was substantial, with 42 of 50 ratings concordant (84%) (Table 2). The remaining 8 discordant ratings (16%) included both adjacent-category disagreements (e.g., 0 vs 1 or 1 vs 2) and two extreme disagreements (0 vs 2). Using linear weights for ordinal scoring, the weighted kappa was 0.77 (95% CI, 0.67–0.87), indicating substantial agreement. Discordance was observed primarily in confounding by indication, selection bias, and prevalent user bias domains, suggesting differences in threshold-level interpretation and application of prespecified scoring rules rather than systematic disagreement.

**Table 2.** Comparison of Bias Assessments Between an AI-Assisted Framework and Expert Evaluation in Simulated Studies.

| Study | CBI* |  | Selection Bias |  | ITB** |  | Disease Latency |  | PUB+ |  |
| --- | --- | --- | --- | --- | --- | --- | --- | --- | --- | --- |
|  | Expert | AI | Expert | AI | Expert | AI | Expert | AI | Expert | AI |
| Study 1 | 0 | 2 | 2 | 2 | 0 | 0 | 0 | 0 | 0 | 2 |
| Study 2 | 2 | 1 | 2 | 2 | 1 | 1 | 0 | 0 | 0 | 0 |
| Study 3 | 2 | 2 | 1 | 1 | 0 | 0 | 2 | 2 | 1 | 1 |
| Study 4 | 0 | 0 | 0 | 1 | 0 | 1 | 0 | 0 | 0 | 0 |
| Study 5 | 2 | 2 | 2 | 2 | 2 | 2 | 2 | 2 | 2 | 2 |
| Study 6 | 1 | 2 | 2 | 2 | 2 | 2 | 2 | 2 | 2 | 2 |
| Study 7 | 2 | 2 | 2 | 2 | 2 | 2 | 1 | 1 | 1 | 0 |
| Study 8 | 0 | 0 | 2 | 2 | 2 | 2 | 2 | 2 | 2 | 2 |
| Study 9 | 0 | 0 | 0 | 0 | 0 | 0 | 0 | 0 | 0 | 0 |
| Study 10 | 0 | 0 | 2 | 2 | 0 | 2 | 0 | 0 | 0 | 0 |

\*= Confounding by indication
\*\*=Immortal time bias
+ =Prevalent user bias
Highlighting indicates discordant scores; shaded cells represent differences between expert and AI platform assessments (Expert = orange, AI platform= pink).

## DISCUSSION

In this validation study of the EpiVise platform we found good agreement between the framework’s assessment of biases and assessments by experts in published studies. Moreover, we found excellent agreement between the framework’s assessment of biases and assessments by experts in the fictitious scenarios. In the second analysis, agreement between expert and EpiVise assessments for immortal time bias was observed in nine of ten simulated studies. Given that immortal time bias is a relatively complex and frequently overlooked source of error in observational research, this finding suggests that EpiVise may be able to reliably identify this bias from study text and could provide useful methodological support when expert review is not readily available.

The tsunami of medical evidence available to patients, clinicians, and policy makers on a daily basis has made it more challenging than ever to tease out ‘signal’ from ‘noise’.^22^ Contradictory findings from high-impact research, such as recent reports linking acetaminophen use to autism risk, can lead to significant unintended consequences and unnecessary public concern. This variability is partly driven by the absence of standardized approaches and inconsistent application of expertise in identifying methodological biases during the peer review process.

Currently, risk-of-bias assessment tools like ROBINS-I exist, and ROBINS-E provides comprehensive frameworks for bias assessment in observational studies.^23,24^ However, their application relies on subjective interpretation of data and substantial expertise in causal inference. This may limit reproducibility, inter-rater agreement, and scalability, particularly in peer review or single-study evaluation. Although many epidemiologic biases can be grouped under broad domains such as confounding, selection bias, and measurement error as captured by ROBINS-I, more nuanced biases such as immortal time bias, may be overlooked when assessments are restricted to these general categories, leading some researchers to use other tools to identify more specific biases, mainly time-related biases, for a more comprehensive assessment.^25^

Of note, immortal time bias can fall under both selection bias and exposure misclassification. Moreover, without explicit definitions and understanding of the bias structure, it may be missed by reviewers who lack formal causal inference training. The framework addresses these issues by applying predefined, rule-based criteria that enable consistent identification of such biases independent of reviewer expertise.

This study has some limitations that warrant consideration. Because no universally accepted gold standard exists for identifying and grading epidemiologic bias in observational studies, we evaluated the framework against expert consensus. This reflects concordance with expert judgment rather than absolute study validity but is consistent with common validation approaches used for risk-of-bias instruments and other expert-coded assessment tools.

Second, although consensus was used, assessment of epidemiologic bias remains partly subjective, particularly for more nuanced biases. Third, the number and range of included studies were relatively limited, which may affect generalizability across different study designs and clinical contexts; however, the included studies still encompassed the key sources of bias most relevant to study validity. Because the framework was developed by the investigators, some degree of alignment with expert ratings cannot be entirely excluded. Finally, important biases beyond the six included in the study, such as measurement error, were not included. Future versions will include a more comprehensive range of biases.

As the volume of observational research continues to grow, scalable approaches to evaluating study validity are increasingly needed. AI-assisted, expert-informed frameworks have the potential to improve the consistency and transparency of bias assessment, helping distinguish true causal signals from spurious findings. Future work should focus on broader validation across study designs and integration into peer review to strengthen the overall reliability of scientific evidence.

## Data Availability

All data referred to in this manuscript are available from the corresponding author upon reasonable request.

